# Sleep Instability in Paradoxical Insomnia is Associated with Perception of Sleep

**DOI:** 10.1101/2022.08.23.22279121

**Authors:** Evelyn Lo, Hsin-Jung Tsai, Albert C. Yang

## Abstract

**Objective:** The main clinical characteristics that diversify paradoxical insomniacs from objective insomnia patients remain unclear. The current study systematically examined the difference in sleep-related, subjective-and objective parameters between insomnia patients with or without misperception.

**Methods:** Patients who reported having poor sleep quality (subjective sleep efficiency; SE ≤ 85%) on both weekends and weekdays and difficulty falling asleep in the Sleep Heart Health Study were examined in this study. Patients with objective SE ≤ 85% and > 85% were then classified as objective insomnia (n = 58) and paradoxical insomnia (n = 61), respectively. The differences in demographic data, subjective sleep quality, daytime function, and objective sleep architecture measured by polysomnography, were assessed between the groups.

**Results:** Though there were no significant group differences in the demographics between objective insomniacs and paradoxical insomniacs, paradoxical insomniacs reported significantly poorer daytime function than patients with objective insomnia. Moreover, paradoxical insomniacs’ subjective sleep evaluation on recording day was significantly higher than habitual reports. The main finding from this research was that the transition indices from stage N3 to stage N2 or N1 was significantly different between the two groups, which could indicate that sleep instability may be a factor in leading paradoxical insomnia patients to underestimate their sleep efficiency.

**Conclusions:** Our findings indicated a possible link between sleep misperception and the microstructure of sleep, specifically the sleep-state instability. The interplay between the neurobiology of sleep instability and perception of sleep needs further investigation.

## 1. Introduction

Paradoxical Insomnia, also known as sleep state misperception, is defined by the International Classification of Sleep Disorders, Second Edition (ICSD-II), as the complaint of severe insomnia to a greater extent than suggested by objective sleep disturbance or daytime function impairment [1]. Though, like all primary insomnias, it has been reduced to a pathophysiological subtype of insomnia disorder in the International Classification of Sleep Disorders, Third Edition (ICSD-III) [2], this sleep disorder is prevalent – depending on the definition used, the prevalence of paradoxical insomnia is within the range of 8% to 66% [3].

No significant evidence of sleep disorder measured by the PSG is present among insomnia patients with sleep state misperception; sleep efficiency (SE) and sleep onset latency (SOL) are all within normal limits [4]. Patients with paradoxical insomnia often underestimate the total sleep time (TST) and overestimate the nighttime SOL [5]. The discrepancy between subjective- and objective sleep quality for paradoxical insomnia is greater than seen in other insomnia subtypes [4].

Paradoxical Insomniacs often report near-constant awareness of the environment (e.g., actively hearing every noise in the house while sleeping in the bedroom) [6], and daytime function impairment reported is often consistent with objective insomnia but is much less severe than expected given the severe level of sleep deprivation reported [7].

Factors that contribute to the discrepancy between self-reported and objective sleep parameters are detailed across numerous researches. Some conclusions were that this phenomenon is independent of age, sex, or laboratory vs ambulatory recording setting, and is especially prominent in insomnia [8]. It is also suggested that a majority of patients with paradoxical insomnia may have misperceived sleep as wake, worry, and brief awakenings [5]. Results from another study indicate that pre-sleep cognitive arousal is negatively associated with sleep misperception [9]. Nevertheless, since limited research has been conducted to evaluate insomnia patients with or without sleep state misperception across a wide variety of subjective and objective sleep structures, little is known about the main factors that diversify paradoxical insomnia patients from other insomniacs.

On the other hand, treatments that tailored this insomnia subtype remain uncertain. One example of treatments aimed to rid paradoxical insomnia patients of this disorder is sleep education, however, only a portion of the subjects responded positively to it - others were unresponsive [10]. Another example of paradoxical insomnia treatments would be Cognitive Behavioral Therapy (CBTi), which proved to decrease the degree of discrepancy between subjective and objective sleep in terms of sleep latency and sleep duration [11], but the extent of improvement decreases with age [12]. These diversifying results encourage further studies to explore the pathophysiology of paradoxical insomnia thoroughly, including using data collected from multi-centered and diverse backgrounds of patients.

The Sleep Heart Health Study (SHHS) provides comprehensive sleep-related data, including subjective- and objective parameters [13,14], which allows the particular examination of specific elements that categorize certain insomniac patients into the paradoxical insomnia group. Thus, the current study adopted this large dataset, since the dataset not only allows the classification of participants into two groups: objective insomnia and paradoxical insomnia, but also provides data on a variety of other variables that can be compared between these two groups, including demographics, subjective sleep quality (i.e., weekday, weekend, or the day after PSG), daytime functions, and objective sleep architecture measured by PSG.

## 2. Method and Materials

### 2.1 Participants

The SHHS, funded by the National Heart, Lung, and Blood Institute, is a multi-cohort study focused on sleep-disordered breathing and cardiovascular outcomes [13,14]. The cohort consisted of 5,804 adults all aged 40 and older and were compiled during two exam cycles, 1995-1998 and 2001-2003; cardiovascular outcomes were tracked until 2010 [13,14].

### 2.2 Objective and Subjective Sleep Quality

The sleep recording was performed by trained and certified technicians with type-II PSG for objective sleep quality assessment [13,14]. The recording montage consisted of an electroencephalogram (EEG), electrooculogram, electromyogram, thoracic and abdominal excursions, airflow, finger-tip pulse oximetry, electrocardiogram, body position, and ambient light status [13,14]. After recording, the standardized visual sleep scoring was then performed by a sleep technologist following the guidelines of American Academy of Sleep Medicine (AASM) to differentiate the sleep stages (i.e., non-rapid eye movement (NREM) sleep stage 1, 2, 3 as N1, N2, N3; rapid eye movement sleep, REM), and arousals in 30-s epochs. The indices for sleep structure were including the total time in bed, TST, SOL (the duration from lights out to the first epoch of any sleep stage), SE (total sleep time/total time in bed; %), wake time after sleep onset (WASO), number of nocturnal awakenings, arousal index, stage transition per hour, and percentage of each stage, calculated as the amount of time spent in each sleep stage divided by total sleep time.

To obtain the subjective sleep quality, self-reported sleep quality and daytime function were evaluated by the questionnaire, including sleep-wake schedules, sleep quality ratings, and the Epworth sleepiness scale [13,14]. For instance,

1. “At what time do you usually FALL ASLEEP on weekdays or your workdays?”
2. “At what time do you usually FALL ASLEEP on weekends or your non-work days?”
3. “How many minutes does it usually take you to fall asleep at bedtime?”
4. “At what time do you usually WAKE UP on weekdays or your work days?”
5. “At what time do you usually WAKE UP on weekends or your non-work days?”
6. “How many hours of sleep do you usually get at night (or your main sleep period) on weekdays or workdays?”
7. “How many hours of sleep do you usually get at night (or your main sleep period) on weekends or your no-work days?

Another morning survey was included in the dataset to examine the subject sleep assessment on the day of PSG recording [13,14], including subjective SOL (S-SOL) and subjective sleep quality (SQ) during the experimental night. Participants’ subjective SE was calculated in two ways. The first method was utilized to calculate weekday-, or workday subjective SE. This was calculated by dividing participants’ reported hours of sleep on weekdays or workdays by the difference between participants’ reported time of falling asleep on weekdays or workdays and participants’ reported time of waking up on weekdays or workdays. The second method of calculating subjective SE calculates the weekend or non-work days subjective SE. The same method was applied to calculate weekend subjective SE. Additionally, we also calculated the subjective SE on the day of PSG measurements by dividing the subjectively reported hours of sleep on the day by the total time in bed (interval between lights out/in-bed time and lights off/out-bed time) on the day of.

### 2.3 Data Enrollment

All the data collected from SHHS-visit 1 (n = 5,804) were initially enrolled. Inclusion and exclusion criteria were determined based on the diagnosis traits of paradoxical insomnia. The current study aimed to identify the differences between sleep insomnia patients with or without sleep state misperception; therefore, participants with evidence of having sleep disorders other than insomnia were firstly excluded. Hence, 2974 participants who had apnea-hypopnea indices (AHI) greater than 4 were identified as sleep apnea patients and thereby removed from this study (n = 2,830).

Secondly, participants who reported habitual subjective SE > 85% were eliminated from the research to meet the criteria of insomnia. The removal of participants with weekend subjective sleep SE > 85% caused 2457 participants to be excluded from the study (n = 373). Likewise, the elimination of the remaining participants with weekday subjective SE > 85% narrowed the participants included in this research to 295 participants (n = 295).

Finally, the insomnia symptom of difficulty falling asleep, as determined by self-reported SOL, was then applied to determine the final group of participants included in this study. After removing 176 participants with self-reported SOL < 30 minutes, 119 participants were identified (n = 119).

### 2.4 Objective Insomnia and Paradoxical Insomnia groups

To evaluate the main differences between sleep insomnia participants with or without sleep state misperception, the 119 participants left in this research were categorized into two groups: objective insomnia and paradoxical insomnia group. They were divided based on objective SE measured by PSG. Those with objective SE > 85% (n = 61) were grouped into the paradoxical insomnia group, and the remaining participants with objective SE ≤ 85% (n = 58) were grouped into the sleep insomnia group.

### 2.5 Statistical Analysis

The demographic data, subjective sleep quality and daytime function, and objective sleep architecture measured by PSG were specifically reported in this study. The chi-squared test (e.g. sex, history of hypertension) or student *t*-test (e.g. SE, WASO) were employed to identify the significant differences between the groups. In addition, paired *t*-test was also employed to examine the differences between habitual and recording subjective sleep evaluation within the two groups. All calculations were done through SPSS ver. 22, and a *p-*value less than 0.05 is considered statistically significant.

## 3. Results

### 3.1 Patient’s demographics by group

Out of 5,804 participants enrolled in the SHHS database, only 119 participants had no evidence of having sleep apnea, reported their subjective SE ≤ 85% on both weekends and weekdays, and reported subjective SOL > 30 min [13,14]. Of the 119 participants, 61 participants had objective SE > 85% and were categorized into the objective insomnia group; 58 participants had objective SE ≤ 85% and were categorized into the paradoxical insomnia group.

As shown in Table 1, there are no statistically significant group differences in age (*t* = 0.93, *p* = 0.36), sex (*p* = 0.27), BMI (*t* = 0.20, *p* = 0.84), ethnicity (*p* = 0.48), education (*p* = 0.78), history of hypertension (*p* = 0.99), diabetes (*p* = 0.12), strokes (*p* = 0.62), or medications status, justified by the intake of sleeping pills (*p* = 0.81), benzodiazepines (*p* = 0.79), and tricyclic antidepressant (*p* = 0.99).

**Table 1.** Participants’ demographics by group.

|  | Objective<br>Insomnia<br>(n = 58) | Paradoxical<br>Insomnia<br>(n = 61) | *p* |
| --- | --- | --- | --- |
| Age (years) | 69.57 ± 10.42 | 67.51 ± 13.73 | 0.36 |
| Male : Female (%) | 45:55 | 34:66 | 0.27 |
| BMI (kg/m<sup>2</sup>) | 28.68 ± 6.40 | 28.46 ± 5.42 | 0.84 |
| Ethnicity (%) |  |  | 0.48 |
| Caucasian | 71 | 79 |  |
| African American | 17 | 10 |  |
| Others | 12 | 11 |  |
| Education (years) |  |  | 0.78 |
| < 10 | 8 | 8 |  |
| 11-15 | 33 | 36 |  |
| 16-20 | 11 | 16 |  |
| 20+ | 0 | 0 |  |
| Medical History (Yes/No) |  |  |  |
| Hypertension | 33/25 | 35/26 | 0.99 |
| Diabetes Mellitus | 9/47 | 3/53 | 0.12 |
| Stroke | 1/56 | 3/55 | 0.62 |
| Medication Status (Yes/No) |  |  |  |
| Sleeping Pills | 8/48 | 6/52 | 0.81 |
| Benzodiazepines | 8/49 | 7/54 | 0.79 |
| Tricyclic Antidepressant | 2/55 | 3/58 | 0.99 |

BMI, body mass index; TCA, Tricyclic antidepressant.

### 3.2 Subjective sleep quality and Daytime Functions

The data in Table 2, specifically those under habitual sleep quality, shows that there are no statistically significant group differences for habitual SOL (*t* = 0.99, *p* = 0.32), habitual SE on weekdays (*t* = -0.17, *p* = 0.86), and habitual SE on weekends (*t* = -0.42, *p* = 0.68).

**Table 2.** Subjective Sleep Quality and Daytime Function.

|  | Objective | Paradoxical |  |
| --- | --- | --- | --- |
|  | Insomnia | Insomnia | *p* |
|  | (n = 58) | (n = 61) |  |
| *Habitual Sleep Quality* |  |  |  |
| Habitual SOL (min) | 50.14 ± 27.09 | 45.79 ± 20.39 | 0.32 |
| Habitual SE on weekdays (%) | 54.91 ± 22.75 | 55.61 ± 21.81 | 0.86 |
| Habitual SE on weekends (%) | 72.03 ± 10.47 | 72.80 ± 9.78 | 0.68 |
| *Sleep Quality after PSG* |  |  |  |
| S-SOL (min) | 45.84 ± 45.32 | 21.02 ± 20.73 | < 0.001 |
| SE recording day (%) | 76.30 ± 22.71 | 90.13 ± 14.90 | < 0.001 |
| SQ (1 to 5) | 2.45 ± 1.12 | 3.12 ± 1.25 | 0.004 |
| Restfulness (1 to 5) | 2.48 ± 1.21 | 3.13 ± 1.26 | 0.01 |
| *Daytime Function* |  |  |  |
| Feel unrested during the day (1 to 5) | 3.16 ± 1.17 | 3.07 ± 1.14 | 0.99 |
| Frequency of not getting enough sleep (1 to 5) | 2.97 ± 1.14 | 3.27 ± 1.16 | 0.65 |
| Frequency of excessive daytime sleepiness (1 to 5) | 2.79 ± 1.15 | 2.77 ± 1.01 | 0.80 |
| ESS | 6.09 ± 4.50 | 7.84 ± 4.42 | 0.04 |

SOL, sleep onset latency; SE, sleep efficiency; PSG, polysomnography; S-SOL, subjective SOL; SQ, subjective sleep quality rated by participant immediately after PSG recording (1 to 5; light to deep); Restfulness, subjective sleep quality rated by participant immediately after PSG recording (1 to 5; restless to restful); ESS, Epworth Sleepiness Scale. Daytime function reported by participants was evaluated with a rating scale for scores 1 to 5 as never to always.

In terms of subjective sleep quality after PSG, there are statistically significant group differences for S-SOL (*t* = 3.60, *p* < 0.001), subjective SE on recording day (*t* = -3.69, *p* < 0.001), SQ (*t* = -2.96, *p* = 0.004), and restfulness (*t* = -2.79, *p* < 0.01).

With regards to daytime function, there are no statistically significant group differences for feeling unrested during the day (*p* = 0.99), frequency of not getting enough sleep (*p* = 0.65), and frequency of excessive daytime sleepiness (*p* = 0.80). However, there is statistically significant group difference for ESS (*t* = -2.09, *p* = 0.04).

Comparing habitual and recording day subjective sleep evaluation within the objective insomnia group, the SE on recording day is significantly different from habitual SE on weekdays (*t* = 5.20, *p* < 0.001), but not significantly different from habitual SE on weekends (*t* = 1.28, *p* = 0.21). Additionally, objective insomniacs’ habitual SOL is not significantly different from S-SOL (*t* = 0.58, *p* = 0.56).

On the other hand, examining paradoxical insomnia group’s habitual and recording day subjective sleep evaluation, paradoxical insomniacs’ SE recording day is significantly different from habitual SE in weekdays (*t* = 10.58, *p* < 0.001), and habitual SE in weekends (*t* = 7.88, *p* < 0.001). Likewise, there is significant difference between habitual SOL and S-SOL (*t* = 7.16, *p* < 0.001).

### 3.3 Objective Sleep Architecture Measured by Polysomnography

As shown in Table 3, the sleep duration section specifically, there is no statistically significant difference between objective insomnia patients and paradoxical insomnia patients for total time in bed (*p* = 0.06) and stage N3 duration (*p =* 0.53), but statistically significant group differences for TST (*t* = -4.60, *p* < 0.001), SOL (*t* = 3.54, *p* < 0.001), stage N1 duration (*t* = 2.34, *p* = 0.02), stage N2 duration (*t* = -3.65, *p* < 0.001) and stage REM duration (*t* = -3.66, *p* < 0.001) were observed.

**Table 3.**
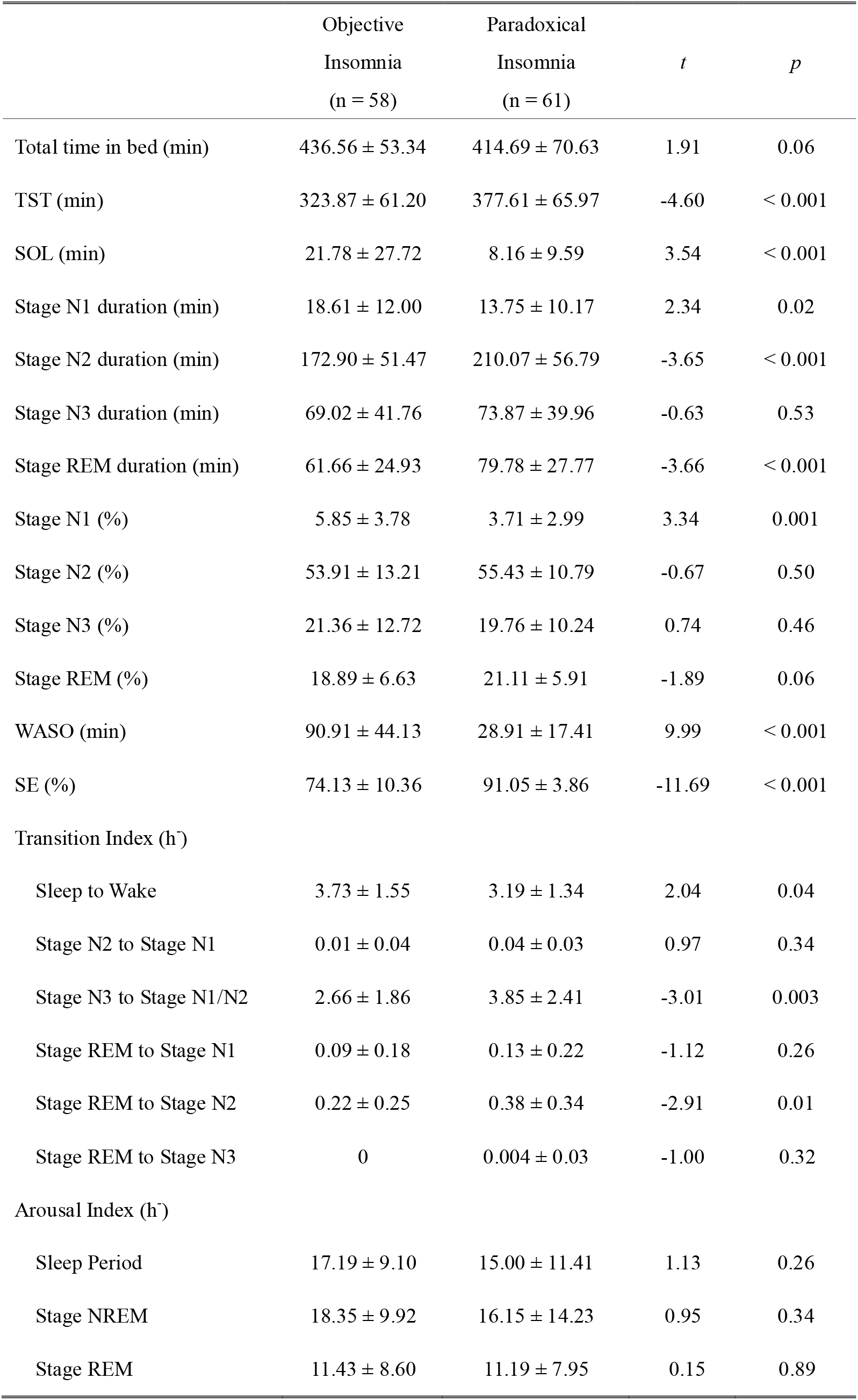
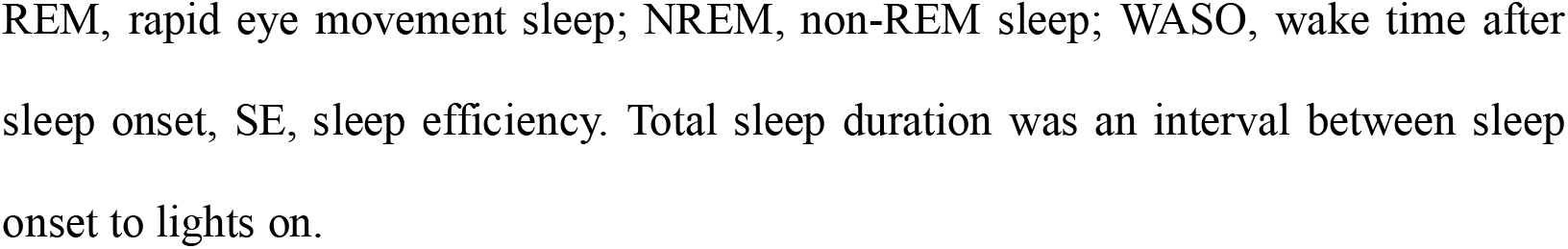
Objective Sleep Architecture Measured by Polysomnography.

|  | Objective<br>Insomnia<br>(n = 58) | Paradoxical<br>Insomnia<br>(n = 61) | *t* | *p* |
| --- | --- | --- | --- | --- |
| Total time in bed (min) | 436.56 ± 53.34 | 414.69 ± 70.63 | 1.91 | 0.06 |
| TST (min) | 323.87 ± 61.20 | 377.61 ± 65.97 | -4.60 | < 0.001 |
| SOL (min) | 21.78 ± 27.72 | 8.16 ± 9.59 | 3.54 | < 0.001 |
| Stage N1 duration (min) | 18.61 ± 12.00 | 13.75 ± 10.17 | 2.34 | 0.02 |
| Stage N2 duration (min) | 172.90 ± 51.47 | 210.07 ± 56.79 | -3.65 | < 0.001 |
| Stage N3 duration (min) | 69.02 ± 41.76 | 73.87 ± 39.96 | -0.63 | 0.53 |
| Stage REM duration (min) | 61.66 ± 24.93 | 79.78 ± 27.77 | -3.66 | < 0.001 |
| Stage N1 (%) | 5.85 ± 3.78 | 3.71 ± 2.99 | 3.34 | 0.001 |
| Stage N2 (%) | 53.91 ± 13.21 | 55.43 ± 10.79 | -0.67 | 0.50 |
| Stage N3 (%) | 21.36 ± 12.72 | 19.76 ± 10.24 | 0.74 | 0.46 |
| Stage REM (%) | 18.89 ± 6.63 | 21.11 ± 5.91 | -1.89 | 0.06 |
| WASO (min) | 90.91 ± 44.13 | 28.91 ± 17.41 | 9.99 | < 0.001 |
| SE (%) | 74.13 ± 10.36 | 91.05 ± 3.86 | -11.69 | < 0.001 |
| Transition Index (h<sup>-1</sup>) |  |  |  |  |
| Sleep to Wake | 3.73 ± 1.55 | 3.19 ± 1.34 | 2.04 | 0.04 |
| Stage N2 to Stage N1 | 0.01 ± 0.04 | 0.04 ± 0.03 | 0.97 | 0.34 |
| Stage N3 to Stage N1/N2 | 2.66 ± 1.86 | 3.85 ± 2.41 | -3.01 | 0.003 |
| Stage REM to Stage N1 | 0.09 ± 0.18 | 0.13 ± 0.22 | -1.12 | 0.26 |
| Stage REM to Stage N2 | 0.22 ± 0.25 | 0.38 ± 0.34 | -2.91 | 0.01 |
| Stage REM to Stage N3 | 0 | 0.004 ± 0.03 | -1.00 | 0.32 |
| Arousal Index (h<sup>-1</sup>) |  |  |  |  |
| Sleep Period | 17.19 ± 9.10 | 15.00 ± 11.41 | 1.13 | 0.26 |
| Stage NREM | 18.35 ± 9.92 | 16.15 ± 14.23 | 0.95 | 0.34 |
| Stage REM | 11.43 ± 8.60 | 11.19 ± 7.95 | 0.15 | 0.89 |

REM, rapid eye movement sleep; NREM, non-REM sleep; WASO, wake time after sleep onset, SE, sleep efficiency. Total sleep duration was an interval between sleep onset to lights on.

In terms of sleep distribution percentage, there are no statistically significant group differences for stage N2 (*p* = 0.50), stage N3 (*p* = 0.46), and stage REM (*p* = 0.06). However, stage N1 (*t* = 3.34, *p* = 0.001) is significantly different between the two groups. In addition, there are statistically significant group differences for WASO (*t* = 9.99, *p* < 0.001), and SE (*t* = -11.69, *p* < 0.001).

Examining the microstructure of sleep, there are statistically significant group differences on some transition indices, including from sleep to wake (*t* = 2.04, *p* = 0.04), stage N3 to stage N1 or N2 (*t* = -3.01, *p* = 0.003), and stage REM to stage N2 (*t* = -2.89, *p* = 0.01), while there are no statistically significant differences from stage N2 to stage N1 (*p* = 0.34), stage REM to stage N1 (*p* = 0.26), and stage REM to stage N3 (*p* = 0.32). Likewise, no statistically significant group difference for arousal index measured in sleep period (*p* = 0.26), stage NREM (*p* = 0.34), or stage REM (*p* = 0.89) was found.

## 4. Discussion

The current study explored the differences in subjective- and objective sleep parameters between insomnia patients with or without sleep-state misperception. Our main findings suggest a possible link between sleep misperception and sleep instability indicated by a significant increase in stage transition from slow-wave sleep to light sleep.

### 4.1 Subjective Sleep Quality and Daytime Function

Although there’s no significant difference in the habitual sleep quality evaluation between the groups, the objective insomnia group’s subjective evaluation of their habitual sleep quality is relatively lower than that of paradoxical insomnia, specifically on SOL, and SE for both weekdays and weekends. Our results showed that the subjectively reported habitual SOL of the objective insomnia group is longer than that of paradoxical insomnia group (objective insomnia vs. paradoxical insomnia; 50.14 ± 27.09 vs. 45.79 ± 20.39, *p* = 0.32). Similarly, the subjectively reported habitual SE on both weekdays and weekends of the objective insomnia group were relatively lower than that the of paradoxical insomnia group, without statistical significance.

Likewise, the subjective sleep quality evaluation after PSG also revealed longer subjective SOL (S-SOL; objective insomnia vs. paradoxical insomnia; 45.84 vs. 21.02 ± 20.73, *p* < 0.001), lower subjective SQ (objective insomnia vs. paradoxical insomnia; 2.45 ± 1.12 vs. 3.12 ± 1.25, *p* = 0.004), lower subjective SE (objective insomnia vs. paradoxical insomnia; 76.30 ± 22.71 vs. 90.13 ± 14.90, *p* < 0.001), and lower restfulness (objective insomnia vs. paradoxical insomnia; 2.48 ± 1.21 vs. 3.13 ± 1.26, *p* = 0.01) among objective insomniacs compared to patients grouped within the paradoxical insomnia group. In summary, both habitual and recording day subjective SQ evaluation are consistent with the trend that objective insomniacs reported worse SQ than that paradoxical insomnia, however, there was no significant group difference for variables that measured habitual sleep quality but there was a significant group difference for variables that measured sleep quality after PSG (*p* < 0.05). Interestingly, by comparing the variables under the after PSG category with the corresponding variables under the habitual sleep quality category, we found a significant increase in the paradoxical insomnia group’s subjective evaluation on the recording day the rom habitual while that of the objective insomnia group generally remained somewhat consistent. Such discrepancies were in line with previous studies that paradoxical insomnia is associated with sleep-related psychological aspects as the cognitive behavioral therapy for insomnia (CBT-i) is more effective among this insomnia subtype than insomnia patients with objective short sleep duration [15,16].

On the other hand, no significant group differences in daytime function between objective- and subjective insomniacs were found, except for the ESS (*p* = 0.04). Patients with paradoxical insomnia reported higher daytime sleepiness than objective insomniacs. Although the scores for both groups were within the normative range [17], our findings suggested that paradoxical insomniacs perceived more sleepiness during daytime activities than insomnia patients with objectively sleep disturbance [4].

### 4.2 Objective Sleep Architecture Measured by Polysomnography

The structures of sleep, as measured by PSG, that display statistically significant group differences are TST, SOL, stage N1 duration, stage N2 duration, stage REM duration, and stage N1%, as shown in Table 3. Without significant differences in total time in bed recorded between the groups, objective insomniacs showed significant shorter TST (objective insomnia vs. paradoxical insomnia; 323.87 ± 61.20 vs. 377.61 ± 65.97, *p* < 0.01), and longer SOL (objective insomnia vs. paradoxical insomnia; 21.78 ± 27.72 vs. 8.16 ± 9.59, *p <* 0.001) than paradoxical insomnia. Moreover, statistically significant differences between the two groups for stage N1% (*p* = 0.001) and WASO (*p* = 0.001) were also observed, indicating that the objective insomnia group spent more time on light sleep stage and longer awake time after sleep onset than paradoxical insomnia. Such statistically significant differences in sleep structure among patients with or without misperception reinforce the fact that the sleep structure of the objective insomnia group, as categorized by subjective sleep evaluation and objective SE < 85%, is worse than that of paradoxical insomnia group [18].

In addition to the macrostructure of sleep, statistically significant group differences for transition indices from Sleep to Wake, stage N3 to stage N1/N2, and stage REM to stage N2 were also observed. The transition index from sleep to wake (objective insomnia vs. paradoxical insomnia; 3.73 ± 1.55 vs. 3.19 ± 1.34, *p* = 0.04) for objective insomnia is statistically significantly higher than that of paradoxical insomnia. This result aligns with the data of WASO and further shows that the sleep stage transition from sleep to wake among objective insomniacs is more frequent than paradoxical insomniacs.

The most important finding that differentiates patients with or without sleep-state misperception objectively is the frequency of stage transition from stage N3 to stage N1/N2 (objective insomnia vs. paradoxical insomnia; 2.66 ± 1.86 vs. 3.85 ± 2.42, *p* = 0.003). This result indicates that the frequency of transitioning from slow-wave sleep to light sleep is greater for paradoxical insomnia patients than that for objective insomnia. One implication that could be drawn based on this result could be that constant transition from deep to light sleep may lead paradoxical insomnia patients to perceive poor sleep quality, and this cannot be identified through the sleep structure examination (i.e., sleep stage duration). Moreover, we found that all transition indices, except for sleep to wake, were lower for objective insomnia than paradoxical insomnia. Transitions to different stages of sleep frequently are associated with sleep-state instability [19], particularly from deep sleep to shallow sleep. Thus, this subtle, sleep-state instability among paradoxical insomnia patients may be a biomarker of this insomnia subtype.

### 4.3 Limitation

While the SHHS provides a wide scope of sleep-related variables, the dataset was conducted focusing on cardiovascular disease and other consequences from sleep-disordered breathing, but it lacked psychological assessments (e.g., depression, psychomotor behavior). To overcome this limitation, we used history of medication intake as a surrogate to evaluate the mental health condition of each participant. Additionally, it is also known that delayed sleep phase disorder (DSPD) is a common disorder in which individuals get to sleep late at night but wake at a normal time in the morning in response to their social clock, resulting in insomnia symptoms and daytime sleepiness [20]. Although there’s no differential diagnosis nor indicators that can be applied in this study to specify if eligible patients were having DSPD, data enrolled in this study all met the subjective SE < 85% on both weekdays and weekends to minimize this potential confounding factor.

## 5. Conclusion

With a wide range of variables provided in SHHS and the immense sample size and diversity of participants, the current study managed to evaluate the significant differences between objective insomnia and paradoxical insomnia. Our findings indicated a significant relationship between sleep-state instability and perception of sleep, and provide a new perspective on the pathophysiology of sleep misperception. The interplay between the neurobiology of sleep instability and perception of sleep needs further investigation.

**Figure 1.**
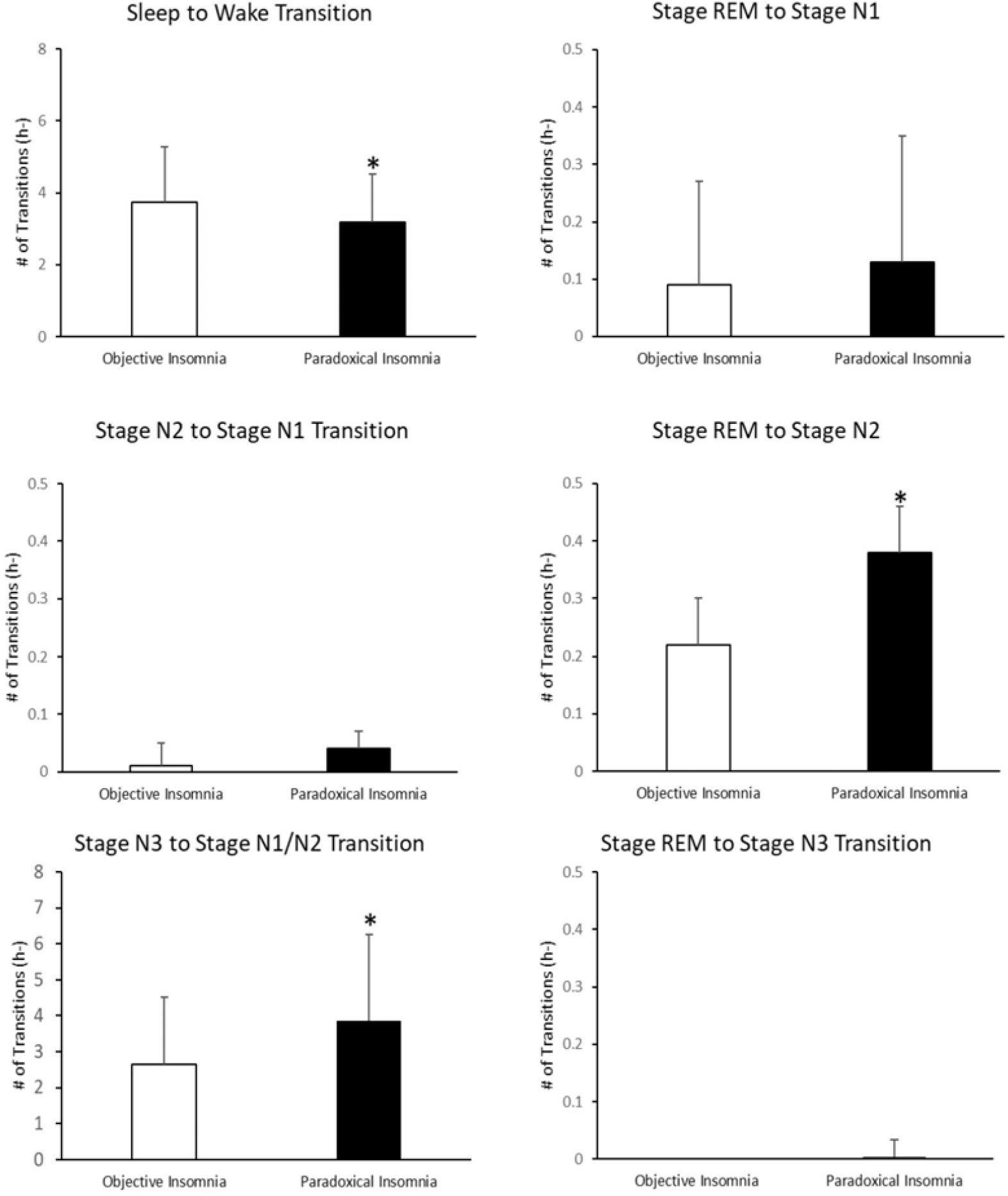
Transition indices from between stages for objective insomnia group and paradoxical insomnia. The transition index from stage N3 to stage N2/N1 of the paradoxical insomnia group is significantly higher than that of objective insomnia group. *, *p* < 0.05, compared to objective insomnia. Data expressed as mean ± standard deviation.

## Data Availability

All data produced in the present work are contained in the manuscript

## Acknowledgement

The Sleep Heart Health Study (SHHS) was supported by National Heart, Lung, and Blood Institute cooperative agreements U01HL53916 (University of California, Davis), U01HL53931 (New York University), U01HL53934 (University of Minnesota), U01HL53937 and U01HL64360 (Johns Hopkins University), U01HL53938 (University of Arizona), U01HL53940 (University of Washington), U01HL53941 (Boston University), and U01HL63463 (Case Western Reserve University). The National Sleep Research Resource was supported by the National Heart, Lung, and Blood Institute (R24 HL114473, 75N92019R002). This study was supported by the Ministry of Science and Technology with grants to Dr. Albert C. Yang [MOST-110-2321-B-A49A-502; MOST-110-2628-B-A49A-509; MOST-110-2634-F-A49-005]. Dr. Albert C. Yang was also supported by Mt. Jade Young Scholarship Award from the Ministry of Education, Taiwan, as well as the Brain Research Center, National Yang Ming Chiao Tung University and the Ministry of Education (Aim for the Top University Plan), Taipei, Taiwan. All the funding resources provided financial supports only and had no other role in study design, data collection, analysis or interpretations.

## Notes

### Competing Interest Statement

The authors have declared no competing interest.

### Funding Statement

This study did not receive any funding

### Author Declarations

National Sleep Research Resource (NSRR) approved.

